# Association between health insurance status and care-seeking behaviors for tuberculosis diagnosis and full treatment among Myanmar migrants in southern Thailand

**DOI:** 10.64898/2025.11.29.25341267

**Authors:** Nyan Lin Htet, Wit Wichaidit

## Abstract

**Introduction:** Myanmar migrant workers in Thailand face elevated tuberculosis (TB) risk due to crowded living conditions, labor-intensive work, and limited access to healthcare. Health insurance may influence care-seeking behavior and treatment completion, yet evidence among this population is limited. The objectives of this study are: 1) to assess the extent to which health insurance status is associated with intended care-seeking behavior for tuberculosis diagnosis among migrants when developing symptoms, and; 2) to assess the extent to which health insurance status is associated with the intention to receive the full course of treatment for tuberculosis in Thailand if given the diagnosis

**Methods:** We conducted a cross-sectional study among 330 Myanmar migrant factory workers using a self-administered questionnaire written in Myanmar language. Key outcomes included intention to seek formal healthcare if TB symptoms developed and intention to complete the full course of TB treatment if diagnosed. We used logistic regression analyses to assess the association between the exposure and outcomes with adjustment for age, sex, income, education, marital status, length of stay in Thailand, and TB history.

**Results:** Most participants reported that they had health insurance (96.1%), and three-quarters (73.3%) reported that they would seek formal healthcare if they developed TB symptoms, and more than four-fifths (83.6%) reported that they would complete the full course of TB treatment in Thailand if diagnosed. Uninsured participants were more likely to not seek formal care (37.5% vs. 17.5%; Adjusted OR = 4.71, 95% CI = 0.60, 36.88). All uninsured respondents who answered the treatment question indicated they would complete the full treatment.

**Conclusion:** Uninsured workers were less likely to seek formal care than insured workers when they developed TB symptoms, but were universally intent on staying in Thailand for the full course of treatment if diagnosed with TB. Limitations regarding the measurement question, selection bias, and information bias should be considered in the interpretation of the study findings.

## INTRODUCTION

Thailand is a major destination hub for migrant workers in South East Asia, with over 4 million registered migrant workers representing 7.5% of the country’s total workforce (ILO, 2025; Rapid Asia Co., Ltd., 2025). Up to 2.3 million registered and 1.8 million unregistered migrant workers in Thailand are Myanmar nationals(IOM Thailand, 2024). Myanmar migrant workers are predominantly in low-wage, labor-intensive industries such as construction, seafood processing, agriculture, and factory manufacturing, which often involve unsafe working conditions, limited safety precautions, and overcrowded environments (Rapid Asia Co., Ltd., 2025). Migrant workers frequently face increased health risks, including from infectious diseases such as tuberculosis (TB), further exacerbated by living in overcrowded dormitories, working irregular hours, and restricted access to health care services (Tschirhart et al., 2016).

Migrants delay TB care-seeking due to limited awareness and concerns about losing jobs or facing deportation ((Homkaew, 2025; Wichai et al., 2012), language barriers, and costs (Wongsuwanphon et al., 2024). Health insurance is a key enabling factor that helps to overcome barriers regarding costs. Insured migrants report feeling less stigma and more secured about their status than undocumented migrants in the absence of other supportive policies (Oo et al., 2022). However, despite Thailand’s formal insurance schemes, e.g., the Migrant Health Insurance and Social Security programs, coverage gaps remain. For example, only 62% of Myanmar migrant workers in Songkhla Province were covered under the social security system (Songkhla Labor Office, 2025; Songkhla SDHSO, 2025). The majority of migrants in southern Thailand rely on self-medication when experiencing TB-like symptoms due to concerns regarding costs and deportation (Naing et al., 2012). Thus, we hypothesize that there are differences regarding care-seeking behavior when developing TB symptoms and intention to complete treatment when diagnosed with TB between Myanmar migrants in Thailand who do and do not have health insurance.

Despite increasing attention to migrant health issue, few studies have explored the extent to which the availability of health insurance is associated with care-seeking and treatment intentions among migrant workers in LMIC context. Such studies can provide basic information that is of interest to policymakers and health system stakeholders in tuberculosis, health services, and migrant health. Thus, the objectives of this study are: 1) to assess the extent to which health insurance status is associated with intended care-seeking behavior for tuberculosis diagnosis among migrants when developing symptoms, and; 2) to assess the extent to which health insurance status is associated with the intention to receive the full course of treatment for tuberculosis in Thailand if given the diagnosis.

## METHODS

### Study Design and Setting

This study employed a cross-sectional design using a structured, self-administered questionnaire. It was conducted among Myanmar migrant factory workers in Songkhla Province, Thailand, focusing on selected factories with a significant proportion of Myanmar workers.

### Population and Sample Size Calculation

The study population consisted of Myanmar migrant factory workers who met the inclusion criteria: (1) aged 18 years or older; (2) currently employed full-time at the participating factories during data collection; and (3) able to read and write in Burmese. Exclusion criteria were: (1) holding Thai citizenship (valid Thai national ID), and (2) currently undergoing tuberculosis (TB) treatment. Participants could withdraw from the study at any stage without consequence to their employment or healthcare access. Each question included a “prefer not to say” option to ensure privacy and minimize discomfort.

Sample size estimation was performed using the n4Studies application for a finite population proportion. We assume a definite population size of N = 47,404 according to data from the Songkhla Provincial Labor Office (Provincial Labour Office Songkhla, n.d.). We also assumed that 6.4% of migrants have TB-suspicious symptoms (n=0.064) (Naing et al., 2012) that we want to present the findings at 3% margin of error (delta=0.03), at 95% level of confidence (z□□ α/2 = 1.96), with a relatively low intra-cluster correlation coefficient (ρ = 0.010). We also assumed and that each factory has at least 30 workers (m=30 workers per factory). Thus, the calculated unadjusted sample size was 254.33, with a design effect of 1.29, resulting in an adjusted sample size of 328.09, which was rounded up to 330 participants.

### Study Instrument

Our study instrument was a Myanmar language paper-based structured self-administered questionnaire comprising six sections: A) Socio-Demography, Migration Information & Health Insurance Status; B) Environmental Risk Factors; C) Occupational Risk Factors; D) Medical Risk Factors; E) Behavioral Risk Factors; and F) Health-Seeking Behaviors for Treatment. We developed the questionnaire in English, translated to Myanmar language, and back-translated to English. We then checked the original and the back-translated versions, identified discrepancies, and made changes to the Myanmar language translation accordingly. We then pilot-tested the questionnaire among a small group of Myanmar migrants and used the feedback to finalize the study instrument.

### Study Variables

#### Health Insurance Status

Health insurance status was assessed by asking migrants whether they had any health insurance. Participants who answered “Yes” were considered insured, while those who answered “No” were considered uninsured. Participants who preferred not to answer were excluded from the main analyses.

#### Intended Care-Seeking Behavior for Tuberculosis Diagnosis

Participants were asked about their intended action if they developed TB symptoms. Those indicating that they would seek care from formal healthcare providers (government hospital, private hospital, or the factory clinic) were classified as intending to seek formal healthcare. We considered those who answered that they would self-medicate (purchasing medical from a local pharmacy, vendor, convenient store, app) or take other actions to report that they would not seek formal healthcare. Responses of “Not sure” or “Prefer not to say” were excluded from the main analyses.

#### Intention to Receive Full Course of TB Treatment

Participants were asked whether they intended to complete a full course of TB treatment if diagnosed in Thailand. Those responding “Yes” were classified as having the intention, and those responding “No” were classified as not having the intention. Responses of “Not sure” or “Prefer not to say” were excluded from the main analyses.

#### Characteristics of the Study Participants

Key socio-demographic and migration characteristics included age, sex, income, education, marital status, length of stay in Thailand, and history of TB diagnosis. These variables were included in Table 1 for descriptive purposes and as potential confounders in adjusted analyses for outcomes 1 and 2.

**Table 1.** Characteristics of the study participants (n=330 migrants)

| Characteristic | Frequency (%),<br>unless otherwise<br>specified |
| --- | --- |
| **Sex** |  |
| Male | 126 (38.2) |
| Female | 202 (61.2) |
| Prefer not to say | 2 (0.6) |
| **Age in years [median (Q1, Q3)] (n=327)** | 30 (24, 37) |
| **Personal monthly income in Thai Bahts (THB)** |  |
| 5,000-9,999 (THB) | 181 (54.8) |
| 10,000-14,999 THB | 107 (32.4) |
| 15,000-19,999 THB | 17 (5.2) |
| 20,000 THB or more | 5 (1.5) |
| Prefer not to say | 20 (6.1) |
| **Education** |  |
| No formal education | 8 (2.4) |
| Attended primary school, did not graduate | 30 (9.1) |
| Graduated from primary school, did not attend middle school | 40 (12.1) |
| Attended middle school, did not graduate | 77 (23.3) |
| Graduated from middle school, did not attend high school | 45 (13.6) |
| Attended high school, did not graduate | 68 (20.6) |
| Graduated high school, no post-secondary education | 29 (8.8) |
| Attended post-secondary education, did not graduate | 18 (5.5) |
| Finished tertiary education | 13 (3.9) |
| Prefer not to say | 2 (0.6) |
| **Marital status** |  |
| Single | 153 (46.4) |
| Married | 139 (42.1) |
| Divorced | 18 (5.5) |
| Widower/Widow | 7 (2.1) |
| Separated | 8 (2.4) |
| Prefer not to say | 5 (1.5) |
| **Duration of stay in Thailand in years [median (Q1, Q3)] (n=299)** | 3 (2, 9) |
| **History of have been diagnosed with tuberculosis** |  |
| Been diagnosed | 19 (5.8) |
| No such history | 300 (91.7) |
| Prefer not to say | 8 (2.4) |
| **Health insurance status** |  |
| Yes | 317 (96.1) |
| No | 9 (2.7) |
| Prefer not to say | 4 (1.2) |
| **Intention to seek care if developed TB symptoms** |  |
| Seek formal healthcare | 242 (73.3) |
| Do not seek formal healthcare | 54 (16.4) |
| Not sure | 23 (7.0) |
| Prefer not to say | 19 (5.8) |
| **Intention to receive the full course of treatment in Thailand if diagnosed with TB** |  |
| Yes | 276 (83.6) |
| No | 12 (3.6) |
| Not sure | 23 (7) |
| Prefer not to say | 19 (5.8) |

### Data Collection

Data collection was conducted between 14 September and 5 October 2025 at selected factories in Songkhla Province. Investigators explained the study purpose in the Myanmar language, provided participant information sheets, and obtained verbal or action-based consent to ensure voluntary and anonymous participation. No personal identifiers were collected. Participants completed the questionnaires within 30 to 45 minutes and submitted them in sealed envelopes or opaque boxes. Each participant received a Myanmar-language TB education leaflet and 50 THB compensation upon completion.

### Data Management

Completed questionnaires were securely stored and later entered into the Kobo Toolbox system. Data were kept on a password-protected server with no personal identifiers. Data quality assurance included completeness checks, numerical coding of categorical responses, and double-entry verification to reduce input errors. The final cleaned dataset was exported to R software for analysis.

### Data Analyses

First, we summarized the characteristics of the study participants using descriptive statistics. Variables included age, sex, income, education, marital status, length of stay in Thailand, history of TB, health insurance status, and intended care-seeking behaviors if they developed TB symptoms. Then, in order to examine the main research questions, we cross-tabulated the health insurance status (having at least one type of health insurance vs. not) and the two outcomes (intention to seek formal care if developed TB symptoms, and intention to complete full TB treatment if diagnosed). We then used unadjusted logistic regression models to calculate crude odds ratios (ORs) with 95% confidence intervals (CIs), then used multivariable logistic regression to calculate adjusted ORs, taking into account other predictors of care-seeking behaviors reported in the literature, including age, sex, income, education, marital status, length of stay in Thailand, and history of TB (Chimbanrai et al., 2008; Helfinstein et al., 2020). We also performed sensitivity analyses on assess the robustness of the study findings. We recoded the responses of “Don’t know” or “Prefer not to say” as having no exposure or no outcome, and re-ran the cross-tabulation and logistic regression analyses. All statistical tests were two-sided, and a p-value of less than 0.05 was considered statistically significant.

### Ethical Considerations

The investigators received ethical approval from the Human Research Ethics Committee, Faculty of Medicine, Prince of Songkla University (REC.68-295-8-2). The Human Research Ethics Committee also approved a waiver of written informed consent and allowed verbal or action-based consent by the participants.

## RESULTS

A total of 330 migrants agreed to participate and answered our study questionnaire (*Table 1*). Most participants were female, earned less than 10,000 THB per month, single, with a middle school education or less. The median age was 30 years, and the median length of stay in Thailand was 3 years. Nearly all (96.1%) reported having health insurance. Nearly three-quarters (73.3%) intended to seek formal healthcare if they developed TB symptoms, and more than four-fifths (83.6%) would receive the full course of treatment in Thailand if diagnosed with TB.

With regard to the association between having health insurance and intended behaviors, migrants without health insurance were more likely than those with health insurance to not seek formal healthcare if they developed TB symptoms (37.5% vs. 17.5%; Adjusted OR = 4.71, 95% CI = 0.60, 36.88) (*Table 2*). None of the migrants without health insurance and a very small proportion of migrants with health insurance answered that they would not stay in Thailand to receive the full course of TB treatment if diagnosed with the disease (0% vs. 4.3%; Adjusted OR = N/A due to perfect prediction) (*Table 3*). Sensitivity analyses for the first scenario (*Table 4*) and the second scenario (*Table 5*) also yielded similar results.

**Table 2.** Insurance status of the migrants and intended care-seeking behavior for tuberculosis diagnosis if developed TB symptoms.

|  | Seek formal healthcare | Do not seek formal healthcare | Crude OR (95% CI) | Adjusted OR (95% CI)* |
| --- | --- | --- | --- | --- |
| **Insurance** | 236 (82.5%) | 50 (17.5%) | 1 (*Reference*) | 1 (*Reference*) |
| **No insurance** | 5 (62.5%) | 3 (37.5%) | 2.83<br>(0.66,12.24) | 4.71<br>(0.60,36.88) |

\*Adjusted for age, sex, income, education, marital status, length of stay in Thailand, and history of TB

**Table 3.** Insurance status of the migrants and intention to receive the full course of treatment for tuberculosis if diagnosed.

|  | Will stay | Will not stay | Crude OR (95% CI) | Adjusted OR (95% CI)* |
| --- | --- | --- | --- | --- |
| **Insurance (n=286)** | 264 (95.7%) | 12 (4.3%) | 1 (*Reference*) | 1 (*Reference*) |
| **No insurance (n=8)** | 8 (100%) | 0 (0%) | N/A | N/A |

\*Adjusted for age, sex, income, education, marital status, length of stay in Thailand, and history of TB

**Table 4.** Sensitivity analysis on the association between having insurance and intended care-seeking behavior for tuberculosis diagnosis if developed TB symptoms.

|  | **Formal medical care** | **Non-formal medical care** | **Crude OR (95% CI)** | **Adjusted OR (95% CI)\*** |
| --- | --- | --- | --- | --- |
| **Insurance (n=321)** | 270 (84.1%) | 51 (15.9%) | 1 (*Reference*) | 1 (*Reference*) |
| **No insurance (9)** | 6 (66.7%) | 3 (33.3%) | 2.65 (0.64,10.93) | 5.39 (0.69,41.94) |

All answers of “Don’t know”, “Not sure”, and “Refused to answer” recoded as non-exposed and non-outcome.
\*Adjusted for age, sex, income, education, marital status, length of stay in Thailand, and history of TB

**Table 5.**
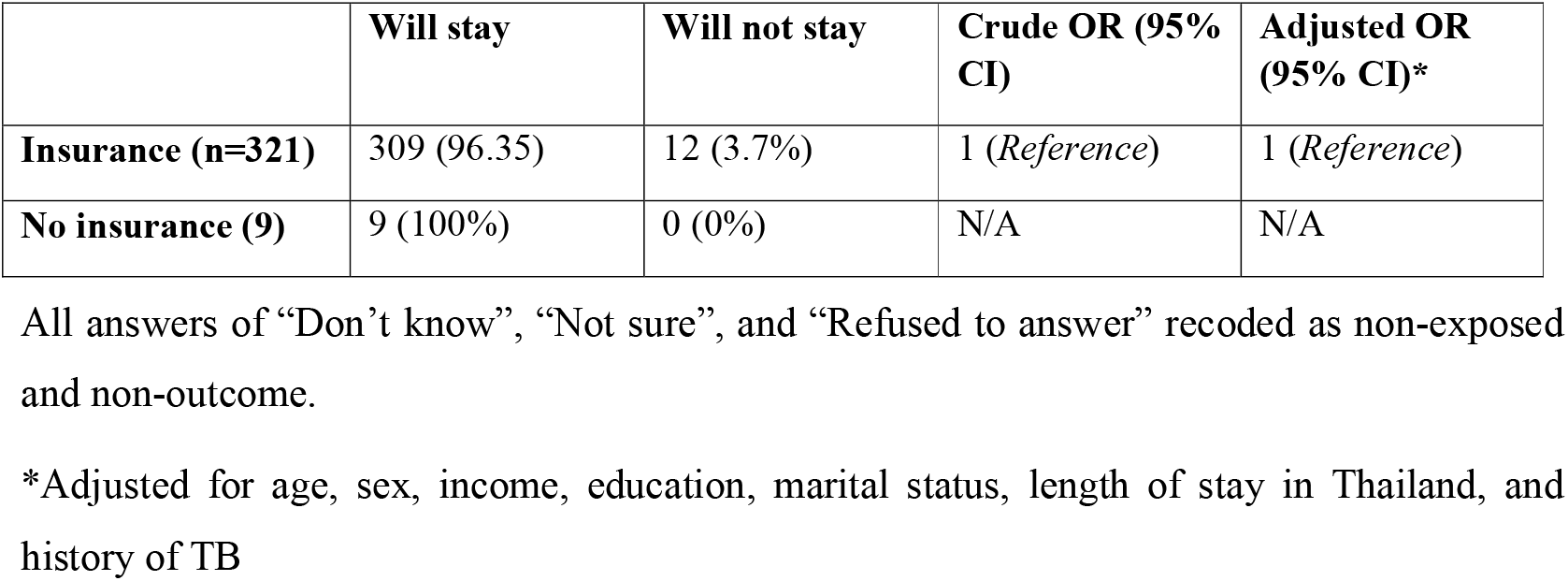
Sensitivity analysis on the insurance status of the migrants and intention to receive a full course of treatment for tuberculosis if diagnosed.

## DISCUSSION

In this workplace-based cross-sectional study, we assessed the association between health insurance status and the intention to seek formal health care for tuberculosis (TB) symptoms and to complete a full course of treatment among Myanmar migrant factory workers in Songkhla province, Thailand. We found that workers without health insurance were less likely to report that they would seek formal healthcare when they developed TB symptoms, but would certainly stay in Thailand for treatment if diagnosed with TB. Our study results are relevant for public health policymakers, TB program managers, and migrant health advocates in designing targeted interventions for this vulnerable population.

The prevalence of not having health insurance in our sample was remarkably low compared to previous reports(Songkhla Labor Office, 2025; Songkhla SDHSO, 2025). The difference was likely a consequence of our study design, which sampled workers from a formal list of factories that were legally required to enroll their employees in the migrant health insurance or social security schemes. The few uninsured individuals in our sample might be new employees awaiting registration. Research conducted in more diverse urban migrant communities in the Bangkok Metropolitan Area shows much lower ownership and use of the Compulsory Migrant Health Insurance scheme (Mon & Xenos, 2015). Thus, the findings of our study may not be generalizable to migrants in less formal sectors such as agriculture, fishing, or domestic work.

Regarding the prevalence of study outcomes, we observed a substantial difference between the intention to seek diagnosis through a formal healthcare provider and the intention to stay in Thailand while receiving treatment. Migrants often self-medicate when having symptoms of illnesses due to treatment costs and fears of deportation (Naing et al., 2012). Even among those with health insurance, issues such as language barriers and fear of losing their job create barriers for care. On the other hand, the seriousness of a confirmed diagnosis may encourage participants to consider staying in Thailand to complete the full course of treatment (Homkaew, 2025; Wichai et al., 2012). The potential influence of social desirability bias, i.e., that participants might have given answers they thought the researchers wanted to hear or that seemed more socially acceptable, could make their reported intentions appear more positive than they actually are.

The association between health insurance and care-seeking behaviors showed a clear trend, with uninsured migrant workers more likely to avoid formal healthcare (Adjusted OR 4.71), which supports the behavioral model of health services use (Andersen, 1995) and is consistent with a previous study showing that insured migrants feel more secure and less stigmatized (Oo et al., 2022). Although the associations were not statistically significant due to the very small number of uninsured participants, the size of the odds ratio indicates a potentially meaningful negative association that should be further explored. Paradoxically, uninsured participants universally stated that they would stay in Thailand to complete treatment if diagnosed with TB. Willingness to stay for treatment may reflect the perceived importance of treatment after diagnosis, or simply an influence of social desirability.

The primary strength of our study was the use of a self-administered questionnaire written in Myanmar language, which might have induced less reactivity or social desirability bias compared to a face-to-face interview. However, a number of limitations should also be considered in the interpretation of the study findings. Firstly, our outcomes were intended behaviors under given circumstances and not behavioral history. Our study findings cannot predict the actual course of action. Secondly, the limited willingness of factories to participate might have introduced selection bias to the study findings. Lastly, self-reported answers from anonymous questionnaires might have nonetheless been influenced by self-serving tendencies or poor memory, potentially introducing selection bias to the study findings.

## CONCLUSION

Our study examined Myanmar migrant factory workers in southern Thailand to see how health insurance affects TB care-seeking and treatment intentions. We observed that nearly all participants had health insurance. We also observed that uninsured workers were less likely to seek formal care than insured workers when they developed TB symptoms, but were universally intent on staying in Thailand for the full course of treatment if diagnosed with TB. Limitations regarding the measurement question, selection bias, and information bias should be considered in the interpretation of the study findings.

## DECLARATIONS

### Data Availability

The dataset and R codes necessary to replicate the findings of this study are available from the corresponding author upon reasonable request.

## Acknowledgments

The authors sincerely thank the Myanmar migrant factory workers who participated in the study for their time and cooperation. We also appreciate the support from factory management teams and community volunteers in Songkhla Province. The authors would like to express their gratitude to Prof. Virasakdi Chongsuvivatwong for his invaluable guidance and inspiration throughout the research process.

